# The effect of strict lock down measures on Covid-19 seroprevalence rate and herd immunity

**DOI:** 10.1101/2020.06.06.20123919

**Authors:** Maher A. Sughayer, Asem Mansour, Abeer Al Nuirat, Lina Souan, Mohammad Ghanem, Mahmoud Siag

## Abstract

**Background:** Covid-19 seroprevalence rates and serological tests are important tools in understanding the epidemiology of the disease and help in the fight against it. Seroprevalence rates vary according to the population studied and the test employed and they range from 0.133 to 25.7%. The purpose of this study is to assess the seroprevalence rate in a population of healthy blood donors living under strict lockdown measures in Jordan which has in total 144 confirmed cases per million population.

**Methods:** Left-over sera and plasma samples from 746 healthy blood donors were tested using a commercially available FDA approved kit having a sensitivity and specificity of 100% and 99.8% respectively. External positive controls were used for validation.

**Results:** More than 80% of the donors were men 18-63 year old and residing in the capital city of Jordan, Amman. All tested specimens were negative yielding a zero seroprevalence rate in this healthy blood donor population.

**Conclusion:** Strict lockdown measures effectively limit intracommunity spread of the infection, however at the cost of lack of any acquired community immunity. Additionally the use of highly specific test is recommended in low prevalence setting.

## Introduction

Since the onset of the Covid-19 pandemic and especially so after the dramatic increase in the incidence of new cases most countries in the world took various measures to control the pandemic. The measures varied from strict lockdown as the case of Hubei, China (1) to more relaxed mitigating policies such as those adopted in Sweden (2). Jordan with a population of almost 10 million adopted strict lockdown measures represented in border closure since mid-March, compulsory supervised quarantine of all people arriving after March 17, 2020 for 2 weeks followed by home quarantine for 2 more weeks, hospital admission with strict quarantine of all newly diagnosed cases regardless of the presence or absence of symptoms, epidemiological case tracing and surveillance with lockdown of neighborhoods, towns and even large districts (governorates) where new cases were detected, prohibiting all kinds of gatherings with closure of mosques and churches, gyms, restaurants and all kinds of shops and imposing various degrees of curfews ranging from complete ones every week end and public holiday to nightly curfew every night since Mid-March till the middle of June when the lockdown measures were gradually relaxed(3). However quarantine measures are still in effect and unnecessary travel still banned with closure of the borders.

The cumulative number of confirmed cases so far (August, 2020) in Jordan is 1438 with only 11 deaths (4). The mode of transmission in Jordan is classified as being clusters of cases (experiencing cases, clustered in time, geographic location and/or by common exposures) as per the WHO (5).

From a relative perspective the number of cases is 144 per million which is less than one twentieth of the world average and among the lowest in the World and the Eastern Mediterranean region (3). It is worth noting that Jordan is among the countries with high testing rates since Jordan performed more than 69,000 tests per million population ranking with the top countries in the world in the number of tests per capita (4, 6).

Seroprevalence data and serological testing are extremely important in combating Covid-19. It fills a gap in the diagnoses and surveillance of previously infected/asymptomatic persons who test negative for the virus (7). It also enhances our understanding of the epidemiological aspects of the disease and helps strategy and decision makers in planning for future actions based on the degree of exposure and community developed immunity or herd immunity.

Several seroprevalence studies were reported and analyzed revealing various seroprevalence rates ranging from 0.133 to 25.7 depending on the population tested. A recent metanalysis concluded that seroprevalence rates are highly region dependent. (8, 9, 10, 11)

The aim of our study is to measure the seroprevalence rate among healthy Jordanian blood donors from the capitol of Jordan, Amman where it is estimated that approximately 40% of the population resides. Additionally we aim to assess the covid-19 asymptomatic infection rate and to indirectly assess the effectiveness of the strict lockdown measures.

## Methods

### Subjects and samples

Left over sera and or plasma collected routinely during the process of blood or apheresis platelet donations were randomly chosen for the study. The donors were healthy asymptomatic subjects between the ages of 18 and 63 who underwent routine screening to determine their acceptability for donation as per standard practice. In addition, starting on the 15^th^ of March additional criteria were adopted which include the requirement of no travel in the 14 days prior to the donation or any contact with a Covid-19 patient or contact with a recent traveler to Jordan. The total number of donations/subjects used for the study was 746 and the donations were distributed over the months of January to June 2020 as follows: 104 in January, 89 in February, 90 in March, 147 in April, 229 in May and 75 in June.

### Testing Methodology

The samples were tested according to the manufactures recommendations using a commercially available FDA approved kit for total immunoglobulins against SARS-Cov-2. The test was performed on Cobas e601 Roche analyzer 6000, using the Elecsys-Anti-SARS-CoV-2 kit (Roche Diagnostics GmbH, Mannheim). This test is a qualitative assay that uses Electrochemiluminescence Method (ECLIA) which is an immunoassay for the in vitro qualitative detection of antibodies (including IgG) to SARS-CoV-2 in human serum and plasma. The assay uses a recombinant protein representing the nucleocapsid (N) antigen for the determination of antibodies against SARS-CoV-2. The test was validated by the manufacturer using 5272 samples including blood donors, diagnostic routine, other corona viruses and common cold panels. The specificity was determined to be 99.81% while the sensitivity was 100% using 40 known positive samples.

Serum samples obtained form 4 previously RT-PCR-confirmed patients who have recovered one month prior to sampling were used as external positive controls.

### Statistics

Descriptive statistics were used to analyze the results.

### Ethical approval

All necessary approvals were obtained.

## Results

Table 1 shows the demographics of the entire group of healthy blood donors enrolled in the study. Over all there were more men than women among the donors tested and most of them were from the capitol Amman of various neighborhoods. Nevertheless a substantial proportion were from other cities and districts in the country.

Table 1

Table 1
| Demographics of the 734 Healthy Donors |  |  |
| --- | --- | --- |
|  | Age | No (%) |
| Males- 626 (85.3%) | 18-30 | 260 (35.4) |
|  | 31-40 | 146 (19.9) |
|  | 41-50 | 66 (9) |
|  | 51-63 | 26 (3.5) |
|  | Unknown | 128 (17.4) |
| Females – 108 (14.7%) | 18-30 | 41 (5.6) |
|  | 31-40 | 12 (1.6) |
|  | 41-50 | 7 (1) |
|  | 51-63 | 4 (0.5) |
|  | Unknown | 44 (6) |
| Residential area Within Amman 597 (81.3%) |  |  |
| Amman, North |  | 194 (26.4) |
| Amman, South |  | 83 (11.3) |
| Amman, West |  | 203 (27.7) |
| Amman, East |  | 117 (15.9) |
| Districts Outside Amman 137 (18.7%) |  |  |
| North |  | 35 (4.8) |
| Central |  | 96 (13.1) |
| South |  | 6 (0.8) |

None of the tested specimens revealed the presence of the antibodies against SARS-CoV-2 indicating a zero percent seroprevalence among this randomly selected group of healthy blood donors. This result indicates no intracommunity transmission is occurring beyond the cases that have already been or are being detected at the entry points to Jordan or their contacts.

## Discussion

To our surprise none of the tested donors was found to be positive for the SARS-Cov-2 antibodies. The method used was validated by the manufacturer and also by our own institution. The test’s sensitivity and specificity as reported by the manufacturer are 100% and 99.81 respectively. The positive predictive value is 96.5% while the negative predictive value (NPV) is 100% at an assumed 5% prevalence. The marketed kits for SARS-CoV-2 antibodies suffer from high false positive rates according to the FDA (12) with a positive predictive value coming down to 27% in some kits at an assumed 5% prevalence rate. Therefore most seroprevalence studies have been criticized for the potential of high false positive rates and that the observed rates are overestimating the real prevalence which is much lower. The kit we used with the given performance characteristics would theoretically detect some false positives but no false negative results (NPV 100%). Since we had not observed any positive result apart from the external controls we did not have to worry about false positive results rather we should have worried about false negative results. But as previously stated the expected false negative rate is 0. Therefore we are confident that we do not have any false negatives and all the negative results are true negative. The seroprevalence rates vary greatly between studies ranging from 0.133 -25.7% depending on the population studied and the method used (10).

The first case of Covid-19 in Jordan was registered on the 2^nd^ of March of this year with more cases registered only 13 days later. However measures to prevent and control the transmission of the infection were implemented as early as February 27^th^ that eventually culminated with the declaration of the state of emergency on the 19^th^ of March and imposed curfew on the 21^st^. Strict lockdown was implemented for almost 2 months. The details of the measures can be found elsewhere (3).

All newly discovered cases regardless of the presence or absence of symptoms or were subjected to quarantined hospitalization and all of their contacts were traced with complete closure for at least 14 days of the area were the cases were identified. These measures are viewed by some as being extreme.

In this study the healthy blood donors’ specimens of sera and plasma were chosen to cover the period from January to June in order to see if the virus was circulating before the reporting of any cases and to serve as a negative control. They also represented various neighborhoods of the capital Amman and even other cities and governorates outside of Amman. They also represented various age groups but males were more heavily represented than females. Although one cannot claim that this is a representative sample of the population of Jordan, nonetheless it may give us a rough idea about the intracommunity transmission of the infection and about the asymptomatic infections.

Our initial surprise can easily then be explained. This seroprevalence rate of zero is probably real despite the limitation of the relatively small sample size. The most likely explanation therefore is the absence of community exposure secondary to the strict lockdown measures undertaken by the authorities in Jordan. The measures implemented were therefore extremely efficient in preventing the disease spread.

On the one hand it really helped the health authorities to have the epidemic under control with no extra constrains or overburden on the health response system with the impressive result of only few hundred cases registered, traced to imported cases form other countries, and of total deaths of less than 10 patients, all of whom were elderly and or with preexisting comorbidities. The number of deaths per million population is extremely low as compared to other countries of the world.

On the other hand Jordan may face a real problem should there be a second wave of infection before the development of an effective vaccine or therapy because of the absence of any community or herd immunity. One may hope before that occurs, a natural decrease in virulence similar to what happened with other corona viruses takes place. This situation of having an immune-naïve community raises concern for some in regards to the strict lockdown measures and weather they were really justified in view of the very low number of cases registered and the early clear evidence that no community spread had occurred. The price to be paid in terms of lack of development of some community immunity may be heavy if a second wave occurs, however proponents of those strict measures argue that the toll in terms of mortality and morbidity could have been much heavier than the country could handle.

Our study is the first one to our knowledge that reveals a zero seroprevalence rate for Covid-19 in healthy blood donors based on the use of a highly specific test. It is also the first and only study from Jordan and the Middle East. Other studies on healthy blood donors revealed low but substantial seroprevalence for covid-19 but none of these studies used the test we used.

In conclusion our study demonstrates that strict lockdown measures are extremely effective in preventing intracommunity spread of Covid-19 and that the use of a highly specific test may help clarify the real prevalence in the community. Whether strict lockdown is necessary or not only time would tell.

## Data Availability

all data are avialble in the manuscript

## Ethical Approval

All ethical approvals were obtained. Consent is waived by the IRB.

